# Suicide- and crisis-risk detection using large language models in mental-health chatbots

**DOI:** 10.64898/2026.01.12.26343914

**Authors:** Samantha Weber, Dustin Klebe, Lukas Wolf, Christopher Aeberli, Stephanie Homan, Nikolas Psathakis, Andrea Casanova, José Garcia Macias, Jack Sykstus, Jie-Ming Li, Barbora Provaznikova, Judith Rohde, Laura Frühschütz, Charlotta Rühlmann, Tobias Welt, Tobias Kowatsch, Birgit Kleim, MULTICAST consortium, Sebastian Olbrich

**Affiliations:** Psychiatric University Hospital Zurich, Department of Adult Psychiatry and Psychotherapy, Psychiatric University Clinic Zurich, 8032 Zürich, Switzerland; Faculty of Medicine, University of Zurich, 8050 Zürich, Switzerland; Institute for Implementation Science in Health Care, University of Zurich, Zurich, Switzerland; Sonia AI (https://soniahealth.com), San Francisco, CA 94104, USA; Department of Psychology, University of Zurich, 8052 Zürich, Switzerland; Healthy Longevity Center, University of Zurich, Zurich, Switzerland; School of Medicine, University of St. Gallen, St. Gallen, Switzerland; Department of Management, Technology, and Economics, ETH Zurich, Zurich, Switzerland

**Keywords:** Suicidality, Emergency Detection, Safety-Critical Systems, Just-In-Time Adaptive Interventions

## Abstract

**Objective:** Large language models (LLMs) are increasingly embedded in mental-health chatbots, yet safe deployment is limited by two unresolved challenges: (1) suicide- and crisis-risk detection lacks a definitive ground truth and is characterized by substantial clinician disagreement, and (2) most evaluations frame risk detection as an offline accuracy task rather than a real-time safety problem. This study aimed to empirically characterize these limitations and to derive design principles for uncertainty-aware, safety-oriented crisis detection in conversational artificial intelligence.

**Methods and Analysis:** We curated a clinician-labeled dataset of 200 real-world conversation segments drawn from a deployed mental-health chatbot. Five clinical experts independently annotated each segment for suicide- and crisis-related risk. Using a single base LLM, we implemented five prompt-defined detection variants with systematically increasing sensitivity thresholds, without task-specific training or fine-tuning. Models were evaluated against clinician consensus labels to quantify false-negative and false-positive trade-offs. Latency analyses assessed feasibility for real-time, per-turn monitoring.

**Results:** As sensitivity increased, the false-negative rate decreased monotonically from 87% to 0%, while false-positive rates rose accordingly. *High*- and *extreme-sensitivity* variants achieved near-perfect (98.9%) and perfect (100%) recall, demonstrating that near–zero–miss crisis detection from natural language is technically feasible in real time (mean latency <1 s). Importantly, model errors aligned closely with cases of clinician disagreement, indicating that misclassifications predominantly reflect irreducible uncertainty rather than model failure.

**Conclusion:** Suicide- and crisis-risk detection in conversational systems is inherently uncertain and should be reframed from an accuracy-oriented classification task toward an online, safety-oriented monitoring problem. Within this framing, near-zero-miss detection is achievable but necessarily incurs elevated false-positives, motivating architectural rather than purely model-level solutions. We propose an operational *emergency mode* in which conservative risk detection operates independently from the conversational model, allowing supportive engagement to be maintained under heightened safety constraints. This layered, uncertainty-aware architecture provides a practical pathway for safer deployment of LLM-based mental-health chatbots without reliance on large training datasets or extensive model optimization.

**What is already known on this topic?:**

- Mental-health chatbots based on large language models are increasingly used for psychological support, but evaluations and real-world incidents show that general-purpose LLMs are unreliable in recognizing suicide- and crisis-related risk and may respond unsafely in high-stakes situations.
- Suicide and crisis risk assessment lacks a stable ground truth, with substantial inter-clinician disagreement even among trained experts, indicating that automated risk detection is inherently uncertain and cannot be treated as a conventional supervised classification task.

**What this study adds:**

- Demonstrates that near-zero-miss suicide- and crisis-risk detection is technically feasible using clinician-validated data and prompt-based sensitivity calibration, without the further need for fine-tuning or large task-specific training datasets.
- Shows that detection errors are largely driven by irreducible uncertainty rather than model failure, as misclassifications systematically align with areas of clinician disagreement, supporting the view that crisis detection is an online monitoring problem rather than a solvable classification task.
- Introduces an architectural safety framework for mental-health chatbots in which conservative, independent risk detection enables an operational *emergency mode* that prioritizes safety while maintaining empathic engagement.

**How this study might afect research, practice or policy:**

- This work reframes suicide- and crisis-risk detection in conversational AI as a safety-oriented, uncertainty-aware problem rather than an accuracy-driven prediction task, challenging prevailing evaluation practices.
- By proposing an architectural separation between risk detection and dialogue generation, it provides a practical, scalable framework for deploying mental-health chatbots that support just-in-time safety interventions without relying on extensive clinical training data.

## Introduction

Mental health conditions affect more than one billion people globally,^1^ and suicide remains a leading cause of premature mortality, claiming between 720,000 and 800,000 lives each year.^2,3^ These numbers reflect a growing gap between individuals experiencing acute psychological distress and the availability of timely, professional support.^4^ Recent advances in large language models (LLMs) have made conversational agents (”chatbots”) widely accessible, offering on-demand psychological support to millions of users through natural, human-like dialogue.^5,6^ Their scalability, anonymity, and low cost make them attractive for those reluctant or unable to seek traditional care. Yet documented incidents in which general-purpose chatbots provided unsafe guidance or failed to recognize acute suicidal intent^7–9^ highlight a critical vulnerability: most LLMs are not designed, aligned, or validated for the detection and management of acute psychological crises. Transparency reports indicate that approximately 0.15% of weekly interactions with commercial chatbots contain signs of suicidal planning, a proportion far too large to ignore at global scale.^10–12^ A central limitation is the difficulty of balancing engagement and safety. Optimizing conversational flow can mask subtle warning signals^13^ such as indirect references to self-harm or hopelessness, whereas overly protective guardrails cause many systems to terminate conversations abruptly after issuing generic hotline instructions.^12,14,15^ Such disconnections reduce legal liability but conflict with well-established principles of crisis intervention, where sustained engagement is often essential for de-escalation.^16^

Despite these challenges, mental-health chatbots remain promising tools for expanding access to psychological support.^15,17^ Addressing this gap would require extensive, clinically validated conversational data to train and evaluate risk-detection models.^18^ However, such data are exceptionally scarce.^19^ Unlike structured clinical datasets, crisis-relevant language emerges unpredictably and does not follow predefined structures. As a result, most prior work on suicide-risk classification relies on synthetic data, post-hoc annotations, or highly curated scenarios, limiting real-world applicability.^19–25^ Moreover, trained clinicians frequently disagree when assessing the same conversational material.^26^ This disagreement reflects not annotation noise, but an irreducible epistemic uncertainty, implying that crisis detection cannot be treated as a conventional supervised classification problem with an attainable optimal decision boundary.

These limitations help explain why existing AI-based risk classifiers, even when technically sophisticated, have struggled to translate into safe real-world deployment. Optimizing models for *offline* accuracy or aggregate performance metrics does not resolve the fundamental challenge of acting under uncertainty in live conversations. In practice, the critical question is not whether suicide risk can be perfectly classified after the fact, but whether emerging danger can be detected early enough to enable timely support. This reframes crisis detection in mental-health chatbots from an accuracy-oriented prediction task toward an *online*, safety-oriented monitoring problem. Rather than inferring a single “correct” label, systems must continuously balance false negatives and false positives in real time, prioritizing safety while tolerating uncertainty. Dedicated risk-detection modules, operating independently of the conversational model, provide a principled way to achieve this separation, allowing empathic engagement to be maintained while safety monitoring is handled conservatively in the background.

Here, we conceptually introduce and validate a LLM–based risk detection system designed for real-world mental-health chatbot conversations. We systematically evaluated five model variants with progressively higher sensitivity thresholds for detecting suicide- and crisis-related language in order to quantify the trade-off between false-negatives and false-positives. This analysis establishes sensitivity levels compatible with safety-critical deployment and motivates the conceptual development of an *emergency mode*, defined as a structured, high-safety operational state that is activated when elevated risk is detected. Crucially, this framework demonstrates how clinically informed risk detection can be implemented without reliance on large, specialized training datasets or extensive model optimization.

## Methods

### Study design and methodological rationale

This study was designed to characterize the inherent trade-offs of crisis-risk detection in conversational mental-health systems, rather than to optimize or benchmark a specific predictive model. Accordingly, we deliberately avoided task-specific model fine-tuning and instead focused on how different sensitivity levels affect false-negative and false-positive behavior under matched conditions. This design allows the results to be interpreted as properties of the detection task and its uncertainty, rather than as performance claims tied to a particular model architecture or fine-tuning.

### Dataset and clinical annotation

We curated a dataset from real conversational segments drawn from users of the mental-health chatbot *Sonia* (https://soniahealth.com). Available in the US, Sonia is a HIPAA-compliant voice-based AI chatbot trained on evidence-based cognitive-behavioural therapy (CBT) and Acceptance and Commitment Therapy (ACT) techniques to provide accessible mental health support through natural conversation, serving thousands of users who often cannot access traditional support systems. Over a three-week period (September 2025), 3,608 user-chatbot exchanges involving 676 clients were screened to identify potentially high-risk moments. From these, we curated 200 segments reflecting the most clinically sensitive situations. Each segment comprised the final ten user-assistant turns preceding an operational safety flag or manual reviewer flag, thereby concentrating cases in which risk was plausible but often ambiguous. The resulting dataset spans 126 distinct clients.

Five clinical experts independently annotated all segments. Before annotation, all clinicians completed standardized onboarding and received detailed written guidelines to promote consistency (Supplementary Materials). Risk assessment was defined with respect to the final user message, using the preceding nine messages as contextual support.

High-risk situations were defined across several major clinical domains. Clinicians were instructed to evaluate whether the final user message expressed any elevated risk within the following categories: 1) suicidal ideation (primary focus, as it represents the most frequently encountered and clinically urgent risk in real-world digital mental health settings), 2) homicidal ideation, 3) psychiatric emergencies, 4) medical emergencies, and 5) abuse or neglect involving vulnerable individuals. Importantly, conversations were not sampled or stratified by category *a priori*. Instead, clinicians applied these categories as an interpretive framework during labeling. Each sample therefore received an overall binary judgment (high vs. low risk) based on clinical assessment across domains, rather than assignment to a predefined class. Thus, the assigned label for each sample refers exclusively to the final user message, with the preceding nine messages providing contextual information to support accurate risk assessment. Individual ratings were then aggregated into a single reference label using the median across the five clinicians, i.e., majority vote.

### Risk Detection Models

All prompt-defined detection variants used the same base LLM (gpt-5-chat-latest, Oct 2025, OpenAI API), the same task description, and the same set of risk categories as provided to clinicians.^27^ This allowed humans and the automated system to operate under matched conceptual definitions without any model-specific training. Aiming at empirically characterizing how model sensitivity affects detection reliability in a safety critical context, we implemented five prompt-defined detection variants with systematically varied sensitivity thresholds. This prompt-only design enabled a controlled comparison of false-negative and false-positive trade-offs across sensitivity settings while keeping the underlying model constant.

To examine how sensitivity calibration affects performance in safety-critical settings, we implemented five prompt variants with systematically increasing sensitivity: *Extreme Low*, *Low*, *Medium*, *High*, and *Extreme High Sensitivity*. Each variant differed along two controlled dimensions:

#### 1. Scope of covered risk

Risk-category definitions and triggering conditions were expanded in a nested manner across models. For example, the *Low Sensitivity* variant required explicit suicidal intent, plan, or access to means, whereas the *Higher Sensitivity* model additionally included passive suicidal ideation (e.g., “I wish I were dead”) and broader cues of psychological danger. Formally, each model’s triggering case set C*_k_* satisfied:

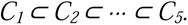

with *k* = 1 denoting *Extreme Low Sensitivity* and *k* = 5 denoting *Extreme High Sensitivity*. This design ensures that higher-sensitivity variants trigger on all cases detected by lower-sensitivity models, plus additional borderline or indirect expressions of risk.

#### 2. Uncertainty policy

Modelprompts also differed in the level of certainty required before classifying a sample as high-risk. We denote the evidentiary threshold for model *k* as *S_k_*:

- *Low Sensitivity* variants require “clear, unambiguous evidence” and instruct “by default, you should not escalate unless unambiguous.”
- *Medium Sensitivity* requires “credible evidence” and instructs “do not escalate unless clear and compelling.”
- *High Sensitivity* require only “indication of danger” or “signs of risk” and instruct “when uncertain about a situation that could involve serious harm, err on the side of caution and escalate.”

This progression establishes a monotonic reduction in the evidentiary threshold:

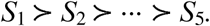

Together, these dimensions approximate a monotonic sensitivity gradient with corresponding activation thresholds *τ_k_*:

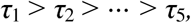

such that more sensitive modelvariants activate on progressively broader sets of risk cues. Although these prompt-variants differ only through natural-language instructions, their behavior empirically resembles a thresholded classifier. That is, each prompt variant can be interpreted as an approximation of a sublevel set

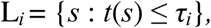

for an implicit underlying risk function *t*(*s*) and threshold τ*_i_*. Increasingly sensitive models therefore expand the set of cues that trigger escalation, providing a practical analogue to sweeping a decision boundary in classical ROC analysis. Importantly, this formulation does not assume the existence of an optimal decision boundary. Instead, it treats sensitivity calibration as a design choice that explicitly trades off missed crises against unnecessary escalation, aligning the evaluation with real-world safety requirements rather than retrospective accuracy. The exact prompts can be provided on reasonable request (see Data Availability Statement).

### Evaluation Framework

To account for stochastic variability in model outputs, each model was run three times on every sample, and the median prediction was used as the final decision (see Supplementary Figure S5). Model predictions were evaluated against the reference clinician labels. Standard classification metrics were computed, including accuracy, precision, recall (sensitivity), specificity, F1score, and the confusion matrix components. However, rather than identifying a single best model(-prompt), the evaluation was intended to illustrate how different sensitivity levels occupy distinct positions along the false-negative-false-positive spectrum, providing empirical grounding for safety-oriented design choices. To visualize this trade-off, we plotted each model’s position in the sensitivity-specificity space using a Receiver Operating Characteristic (ROC) curve.

### System latency measurement

Latency was evaluated to assess feasibility for continous, per-turn monitoring in a hypothetical live conversational system. Thus, we measured end-to-end response latency across 3,000 inference trials. Each trial corresponded to a single user message submitted to the risk detection system. Latency was defined as the elapsed time between submission of a message and return of the corresponding risk classification. Measurements thus reflect model inference time and internal system overhead, but exclude user typing time, client-side delays, and network latency.

### Error Analysis

To better characterize the sources of model error, we conducted a systematic error analysis across all five sensitivity variants. For each model, we identified all samples where its prediction diverged from the reference label (defined by majority vote across the five clinicians of at least three out of five clinicians agreeing on a label). Because the consensus label could still reflect disagreement among up to two annotators, we quantified human uncertainty by counting how many clinicians disagreed with the majority label for each sample (0, 1, or 2). This measure provides a simple index of annotation ambiguity and allows us to contextualize prediction errors relative to underlying human disagreement.

### Patient and Public Involvement

There was no direct involvement from patients in the design of this study. The results are nevertheless intended to inform the development of safer digital mental-health interventions by providing design principles for risk detection that do not require large, specialized training databases or extensive model optimization.

## Results

### Dataset and Clinical Annotations

We curated 200 real-world conversational segments from a total of 3,608 conversations between users and the mental-health chatbot *Sonia*, each independently labeled by five clinical experts. Based on the majority vote, 108 samples (54%) were classified as low-risk and 92 (46%) as highrisk. Interrater agreement reached a Fleiss’ κ of 0.34, indicating fair but variable agreement. Supplementary Figure S3 shows the distribution of votes across annotators.

### Risk Model Performance across varying Sensitivity Levels

We evaluated five prompt-defined risk detection variants: *Extreme Low*, *Low*, *Medium*, *High*, and *Extreme High Sensitivity*, each representing a progressively lower threshold for classifying content as high-risk. Across sensitivity levels, recall of critical content increased from 13% for the *Extreme Low Sensitivity* variant to 100% for the *Extreme High Sensitivity* variant (Table 1). Precision decreased correspondingly from 91.2% to 63.4%, while F1-scores ranged from 0.23 to 0.80, with the *High Sensitivity* model achieving the highest F1-score. Overall accuracy varied between 0.59 and 0.82, with the *Medium Sensitivity* model achieving the highest overall accuracy. In summary, across all model variants, increases in recall (i.e., reducing the number of false-negatives) were consistently accompanied by increases in false-positives, reflecting a systematic shift between false-negative and false-positive errors as sensitivity was increased. Supplementary Figure 1 depicts the confusion matrices for the individual sensitivity levels. Supplementary Fig. S4 shows the performance metrics by model-prompt variant.

**Table 1:** Confusion matrix counts and performance metrics of the risk detection models at different sensitivity levels. Recall is defined as 1−False Negative Rate and False Positive Rate as FP*/*(FP + TN). Bold numbers highlight the best results. *Abbreviations*: TP = True Positives, FP = False Positives, FN = False Negatives, TN = True Negatives, FPR = False Positive Rate

| Model Prompt Variant | TP | FP | FN | TN | Accuracy | F1 | Precision | Recall | FPR |
| --- | --- | --- | --- | --- | --- | --- | --- | --- | --- |
| *Extreme Low Sensitivity* | 12 | 2 | 80 | 106 | 0.590 | 0.226 | 0.857 | 0.130 | **0.018** |
| *Low Sensitivity* | 52 | 5 | 40 | 103 | 0.775 | 0.698 | **0.912** | 0.565 | 0.046 |
| *Medium Sensitivity* | 70 | 15 | 22 | 93 | **0.815** | 0.791 | 0.824 | 0.761 | 0.139 |
| *High Sensitivity* | 91 | 44 | 1 | 64 | 0.775 | **0.802** | 0.674 | 0.989 | 0.407 |
| *Extreme High Sensitivity* | 92 | 53 | **0** | 55 | 0.735 | 0.776 | 0.634 | **1.000** | 0.491 |

The Receiver Operating Characteristic curve (Figure 1) yielded an AUC of 0.90, indicating consistent discriminative ability across threshold settings. Together, these results show that adjusting sensitivity thresholds produces the expected trade-off between minimizing false-negatives and tolerating increased false-positives (Figure 2).

**Figure 1.**
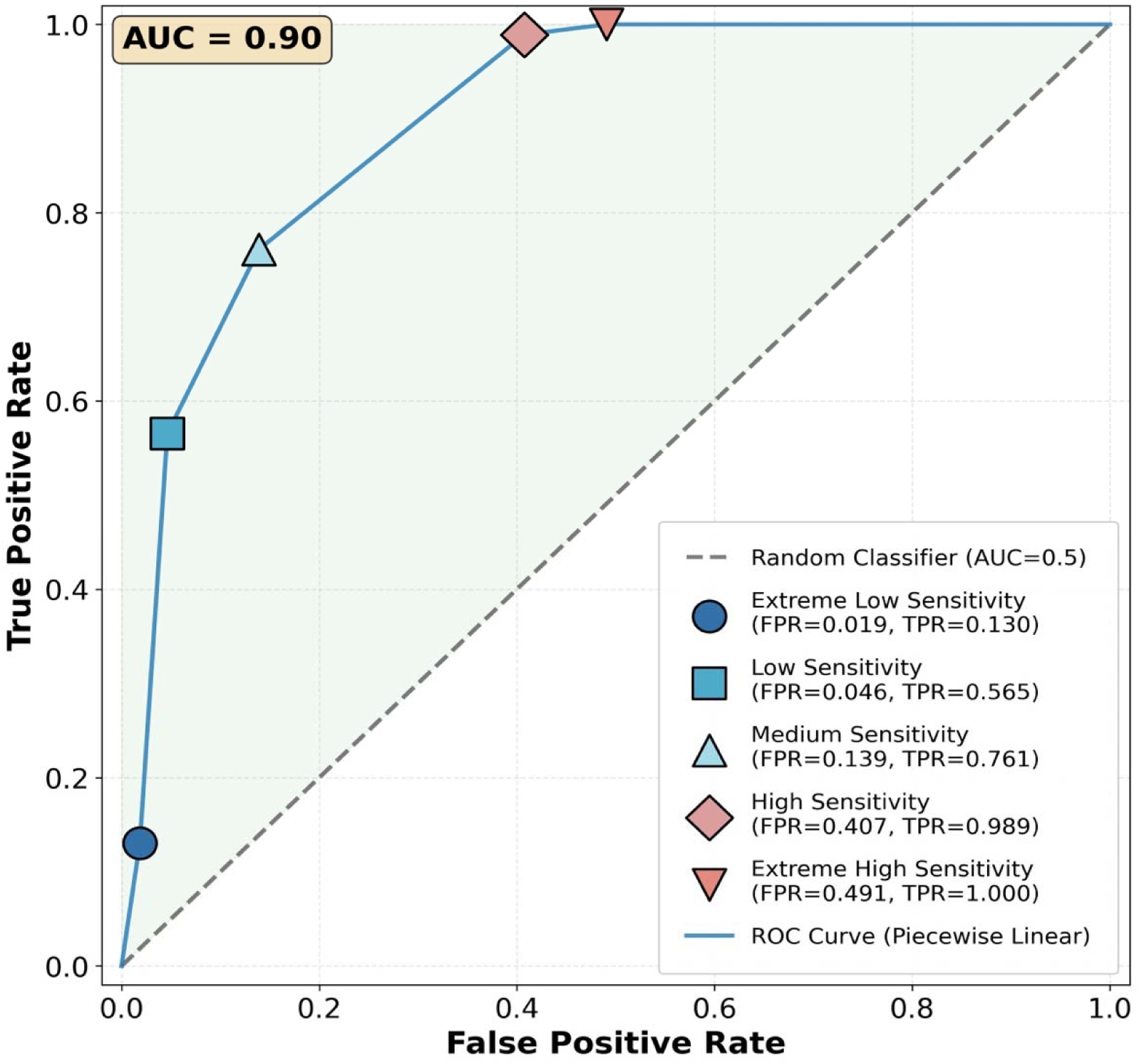
Receiver Operating Characteristic (ROC) Curve Across Sensitivity Levels. The ROC curve compares model variants ranging from *Extreme Low* to *Extreme High Sensitivity*, illustrating the trade-off between true positive rate (TPR) and false positive rate (FPR). The curve, approximated by piecewise linear interpolation, achieved an area under the curve (AUC) of 0.90, indicating excellent overall discriminative performance across sensitivity thresholds. The dataset contains 200 samples with 92 high-risk (positive) and 108 low-risk (negative) cases.

**Figure 2.**
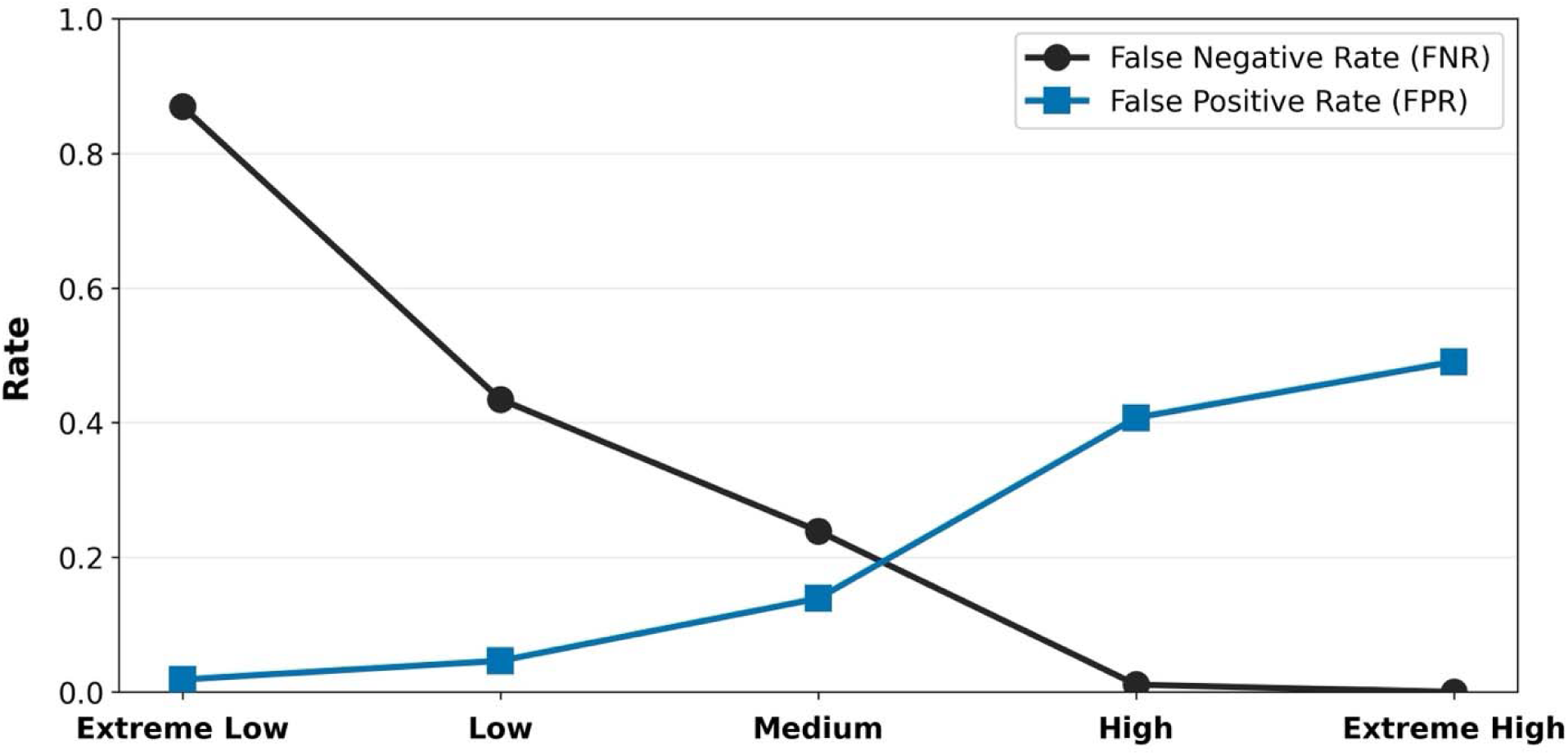
False Negative and False Positive Rates Across the Five Sensitivity Levels. Comparison of false negative (FNR) and false positive (FPR) rates across five model configurations (*Extreme Low -*, *Low -*, *Medium -*, *High -*, and *Extreme High Sensitivity*). As sensitivity increases, FNR steadily decreases, reaching zero in the *Extreme High Sensitivity* model, corresponding to a recall of 1.0 which indicates perfect detection of all high-risk cases. This improvement comes at the cost of increased FPR, reflecting more frequent false alarms. These results highlight the inherent trade-off between minimizing missed emergencies and maintaining specificity in safety-critical risk detection systems.

### System latency measurement

To evaluate real-time feasibility, we measured response latency across 3,000 trials. The distribution showed a primary concentration between 0.5 – 0.8sec (mean = 0.64sec, SD = 0.29sec; median = 0.60sec), with infrequent outliers exceeding 2sec, indicating stable but occasionally variable performance (see Supplementary Fig. S6).

### Error Analysis

As the consensus clinician label represented the agreement of three or more clinicians (majority out of five), disagreement was limited to at most two raters per sample. With that, model errors closely mirrored areas of clinician disagreement. Misclassifications increased systematically as clinician disagreement rose from 0 to 2 annotators, suggesting that ambiguity, rather than model failure, drives many errors. For the *High Sensitivity* variant, only nine errors occurred in unanimously rated cases, compared with 12 when one annotator out of five disagreed and 24 when two disagreed. A similar pattern appeared in the *Extreme High Sensitivity* variant: eliminating false-negatives increased false-positives from 10 (unanimous cases) to 30 (two-disagreement cases). In many such instances, the model’s prediction aligned with one or two clinicians rather than the majority vote, reinforcing that disagreements often reflect inherent uncertainty rather than systematic misclassification. Figure 3 visualizes model performance across the five sensitivity thresholds of the models as a function of interrater disagreement.

**Figure 3.**
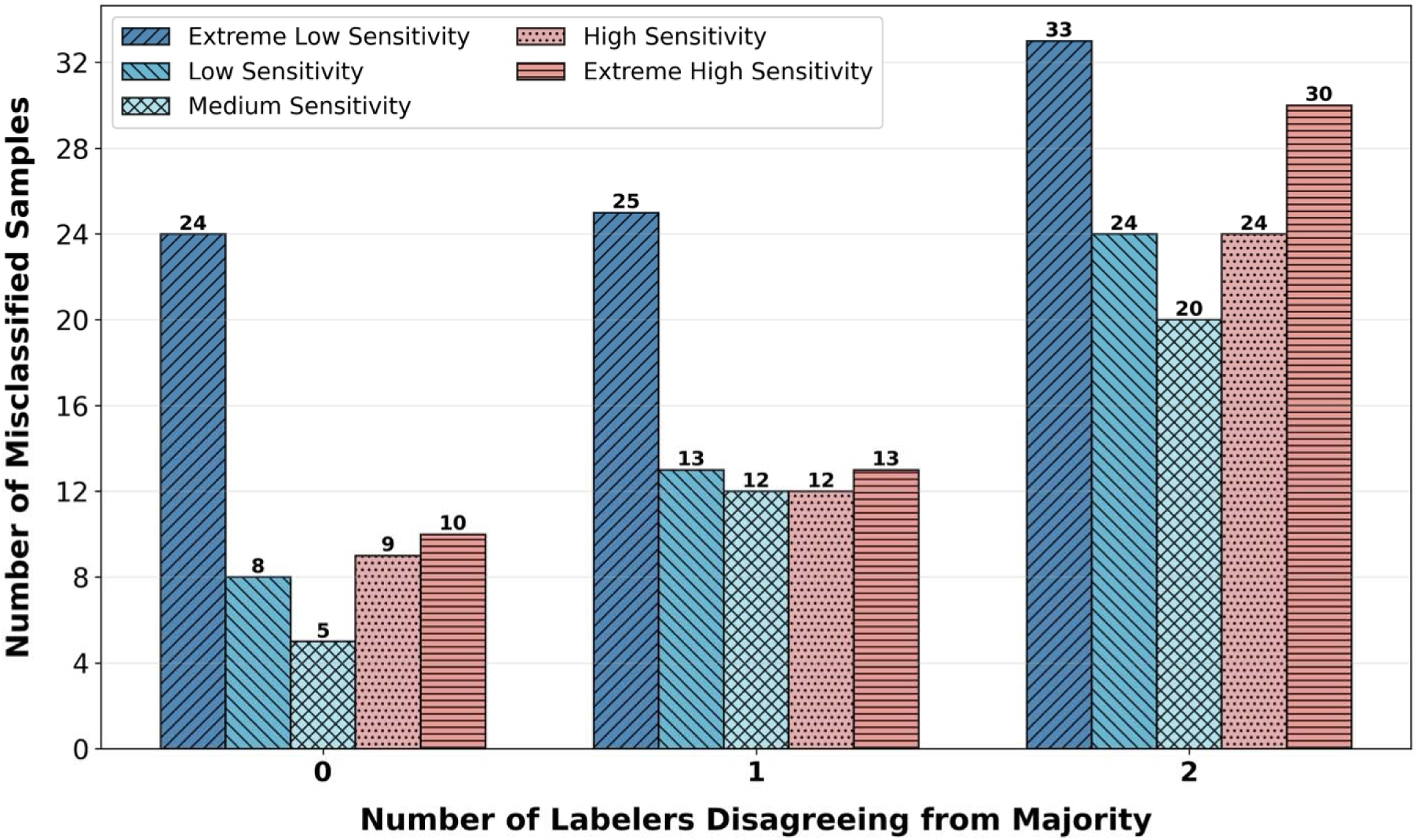
Error Analysis by Labeler Disagreement. Comparison of error counts across five model configurations (*Extreme Low -*, *Low -*, *Medium -*, *High -*, and *Extreme High Sensitivity*), grouped by the number of raters disagreeing from the majority vote. Each group represents the number of misclassified samples where 0 (complete agreement among raters), 1, or 2 rater diverged from the consensus. The results show that error rates increase with greater disagreement among clinical experts, indicating that model misclassifications are more likely in cases where human raters also show uncertainty.

## Discussion

In this study, we evaluated five LLM–based detection variants through prompt-based sensitivity calibration using a clinician-validated dataset of 200 real-world conversation segments labeled for suicide- and crisis-related risk. As sensitivity increased, the false-negative rate (FNR) decreased monotonically, moving from an 87% in the *Extreme Low Sensitivity* variant to 0% (i.e., no missed high-risk cases) in the *Extreme High Sensitivity* variant. In other words, the *High Sensitivity* and *Extreme High Sensitivity* variants achieved near-perfect (98.9%) and perfect (100%) recall while maintaining meaningful discrimination to false-positive cases, demonstrating that LLMs can match clinician consensus on the most safety-relevant classifications. This monotonic relationship reflects the core safety tension in digital mental-health systems: minimizing false-negatives necessarily increases false-positives. A closer inspection of false-negatives and false-positives illustrates how steep this trade-off becomes: The *High Sensitivity* variant missed only one high-risk case (1/92; 1.1%) while producing a 40.7% FPR. Eliminating this final missed case in the *Extreme High Sensitivity* variant required increasing the FPR to 49.1%. Thus, preventing the last ∼1% of false-negatives required accepting an additional ∼8% false-positives. This false-negative–false-positive gradient must also be interpreted in light of human variability: many cases that the model “misclassified” were the same cases on which clinicians themselves disagreed, underscoring that the ground truth in crisis detection is indeed probabilistic rather than absolute. In addition to a near-perfect classification, response-latency analyses further showed that the system operated well within real-time constraints (mean *<* 1s), supporting that risk detection could be continuously executed in real-world conversational turns, which would allow for real-time monitoring within AI-powered chatbot dialogues. Together, these findings provide evidence that LLM-based risk detection can achieve clinician-level identification of high-risk situations in real time, even in conversational segments where clinicians themselves show substantial disagreement. These findings suggest that given a clinician-evaluated system instruction, LLM-chatbots can be safely prompted for crisis- and suicide risk detection within a low-cost environment where no further training data or fine-tuning might be required.

Beyond model-level findings, these results align with and extend growing evidence from broader safety evaluations. McBain *et al.*^20^ and Lauderdale *et al.*^28^ both reported that ChatGPT, Claude, Gemini, and similar models show empathic tone but inconsistent risk judgments, frequently underestimating severity or overescalating ambiguous cases. Furthermore, it was demonstrated that, although chatbot responses to suicide-related queries have become more empathetic and comprehensive over time, critical safety lapses persist, underscoring the unpredictability of unsupervised systems.^14^ Complementary progress has been shown by Balt *et al.*^29^, whose few-shot LLaMA model achieved substantial agreement with experts in coding post-suicide psychosocial-autopsy interviews with relatives. Their findings parallel ours in showing that false-positive-tolerant approaches yield the most reliable outcomes in emotionally complex data. Further conceptual work similarly emphasizes that generative AI can support suicide prevention only when transparency, ethical oversight, and contextual calibration are ensured.^30^ Our results further extend this vision empirically by quantifying the false-negative-false-positive trade-off.

Over the past decade, conversational agents for mental-health have evolved from scripted dialogue systems to LLMs, yet persistent safety and reliability challenges remain. Early systems demonstrated modest improvements in depressive and anxiety symptoms but offered limited contextual understanding and virtually no safeguards for suicidal ideation or crisis management.^31^ Broader evidence from umbrella and scoping reviews confirmed that most available chatbots remain rule-based with few demonstrating robust crisis-handling capabilities.^32^ Early clinical pilots integrating conversational AI into cognitive-behavioral interventions suggested potential for improved engagement and continuity between sessions, yet these implementations relied on constant human supervision and lacked validated risk-detection mechanisms.^33^ Using generative models showed progress in empathy and user acceptance but continues to expose the fragility of current safety mechanisms. Studies in inpatient and outpatient settings indicated that ChatGPT-assisted interactions can enhance perceived support but remain ethically limited by the absence of validated safeguards and transparent escalation pathways.^34,35^ Recent analyses further reinforce these concerns, showing that while LLMs can achieve clinician-level accuracy on well-structured clinical tasks, they remain vulnerable to hallucinations, uncertainty miscalibration, and unpredictable safety behavior under emotionally loaded content or when conversational ambiguity is present.^38–40^ Collectively, this previous work argues that LLMs require structured oversight mechanisms and task-specific guardrails to function safely in high-stakes healthcare environments. Here, consensus guidelines highlighted transparency, informed consent, human oversight, and structured crisis protocols as non-negotiable design principles.^42^ Together, these developments point toward an emerging consensus: mental-health chatbots must operate in real time, continuously adapting to evolving conversational and emotional cues. This trajectory directly aligns with the principles of Just-in-Time Adaptive Interventions (JITAIs^43,44^), which emphasize timely, context-aware responses to user needs. Our study builds on this foundation by embedding threshold-controlled risk detection that does not require excessive training nor large databases. Through prompt-based calibrated sensitivity, it introduces the safeguards identified as missing in prior chatbot research and provides a practical, real-world pathway for safe deployment of LLM-based mental-health systems. Future work may also examine whether lightweight personalization (e.g., based on a user’s typical conversational style or responses to a brief onboarding questionnaire) can improve the balance between false-positive and false-negative without compromising safety.

The present work demonstrates that LLM-based risk detection can operate in real time and achieve near–zero–miss rates under clinically validated conditions. A natural next step is to consider how such detection modules can be integrated into full conversational systems. Because the risk detector in our study functions independently from any response-generation model, it can be optimized exclusively for sensitivity to rare signals without compromising therapeutic quality - a separation that addresses the limitations of current chatbots. Models deployed for emotional support must balance empathy, engagement, safety, and conversational coherence, what might inadvertently suppress safety cues.^14,45^ When detection is handled by a dedicated module, these competing objectives must not interfere.

A second challenge concerns the behavior of chatbots once risk is detected. Many existing systems respond to crisis-related language by issuing generic hotline instructions and abruptly terminating the session.^12,14,15^ Although legally conservative, premature termination can be clinically counterproductive, especially when the user is reaching out at a moment of heightened vulnerability. Along that line, Zortea *et al.*^46^ emphasizes that suicide-risk prediction is inherently uncertain and that prevention depends less on accurate forecasting than on empathic engagement, safety planning, and follow-up care. Likewise, evidence from research on crisis hotlines and chat services underscores the importance of maintaining contact: sustained engagement, clarification of intent, and collaborative safety planning are consistently associated with de-escalation and reduced suicidal ideation.^16,47,48^ Effective crisis work also relies on gathering clarifying information (e.g., the person’s location, the presence of others, or access to means), which can later guide referral to appropriate human support. These principles of non-abandonment, empathic clarification, continuity, and information gathering provide practical guidance for digital systems, even if full human crisis intervention cannot be replicated.

Drawing on this evidence, we propose that future digital mental-health systems may benefit from an operational *emergency mode*: a constrained, safety-focused state that activates when the risk detector identifies elevated concern. Unlike full crisis intervention, *emergency mode* is designed to manage uncertainty and the high FPRs that arise unavoidably in rare-event detection. In this state, the conversational model would operate under more restrictive safety prompts, including 1) prioritize brief clarification of ambiguous cues, 2) provide validated human crisis resources when appropriate, and 3) escalate or terminate only in the most severe cases . This approach would allow conservative thresholds to be used without triggering unnecessary conversation termination or user distress, alternatively even as a long-term vision providing continuous support. This architectural separation mirrors a long-recognized feature of psychotherapy: clinicians engage empathically in the conversation while simultaneously monitoring for subtle indicators of risk in the back of their heads. This parallel workflow of continuous safety appraisal alongside supportive dialogue, is one of the elements that distinguishes therapeutic interactions from ordinary conversation. While digital systems cannot replicate the nuance of human clinical judgment, structuring risk detection as an independent, always-on process allows chatbot systems to approximate this dual-process logic: maintaining user-centered engagement while remaining vigilant to cues requiring heightened safety precautions. Nevertheless, the feasibility of such architectures is strengthened by our latency findings, which show that the risk detector operates well below one second per interaction. This establishes the technical viability of executing continuous risk assessment within live chatbot dialogues without perceptible delay. While our current system evaluates each conversational turn independently, JITAIs^43,44^ provide a broader conceptual framework in which future systems may integrate temporal dynamics across multiple turns, sessions, or days. Taken together, our detection module provides a building block for future JITAI-consistent suicide-prevention strategies in digital mental-health care. Figure 4 illustrates how the conversational model and the backgorund safety system operate in parallel, with the risk-detection module continuously monitoring user input and activating *emergency mode* when safety thresholds are exceeded.

**Figure 4.**
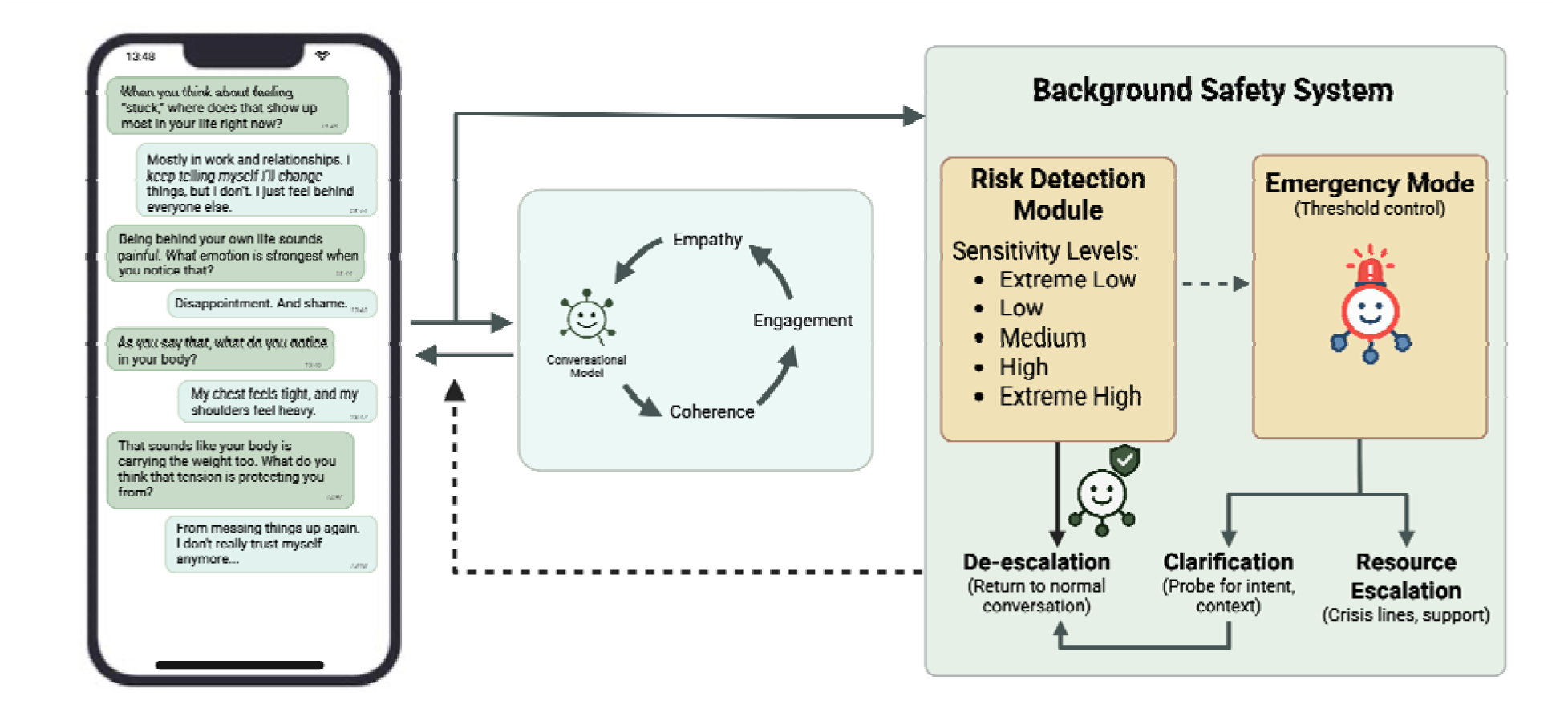
Architectural overview of the conversational model and background safety system. A conversational large language model (LLM) engages the user through empathy, coherence, and therapeutic dialogue (left). In parallel, a background safety system continuously processes each conversational turn. The risk-detection module evaluates incoming text at predefined sensitivity levels (*Extreme Low* to *Extreme High*). When the model detects cues that exceed the threshold, control transitions to *emergency mode*, which operates under stricter safety constraints. Emergency mode conducts brief clarification, provides validated crisis resources when appropriate, and escalates only in severe cases, whereas low-risk signals result in de-escalation back to standard conversation. This separation enables continuous real-time safety monitoring without disrupting supportive dialogue. Figure has been created with biorender.com.

This study has several limitations that should be considered when interpreting the findings. Our evaluation dataset comprises 200 conversational segments, a small to medium-sized sample that requires validation on larger and more diverse datasets. Moreover, we intentionally oversampled high-risk cases (46% of the dataset) to ensure sufficient power for evaluation, which however introduces a prevalence bias. Considering the full corpus of 3,608 user–chatbot interactions, only 92 cases (2.55%) were ultimately labeled as high-risk by clinicians, despite 200 (5.5%) initially being flagged by the operational system. This prevalence is substantially higher than that reported for general-purpose LLMs (∼0.15% according to^11,12^), as expected in a mental-health context such as *Sonia*, yet still represents a rare-event regime. Therefore, while the FPR observed here likely overestimated the rate of false-positives that would occur under natural conditions, the absolute number of false-positives in practice will nevertheless greatly exceed the number of true-positives. This imbalance is a mathematical property of rare-event detection and is not reflected in our deliberately balanced evaluation dataset. A further limitation is the level of interrater agreement. Despite independent ratings by five clinical experts, Fleiss’ κ was 0.34, indicating only fair agreement among clinicians. This variability, however, aligns with prior findings in psychiatric suicide risk assessment, where even trained clinicians show only modest reliability when rating identical cases.^26^ Subtle or indirect expressions of distress often complicate recognition, a challenge well-documented in suicide-prevention research.^24,44^ Suicide risk itself is inherently dynamic and probabilistic rather than categorical, and even structured clinical tools show limited predictive validity.^49^ Furthermore, our evaluation of five variants confirmed a fundamental tension in automated risk detection: reducing false-negatives inevitably increases false-positives. This mirrors long-standing dilemmas in suicide prevention, where interventions maximizing sensitivity can compromise specificity and produce unintended effects such as unnecessary hospitalization or loss of trust^50^, or - in case of chatbots - leaving users to feel misunderstood or dismissed.^16,48,51^ While our proposed architecture mitigates this by pairing conservative risk detection with an emergency mode for appropriate handling of FP, the underlying trade-off remains conceptual rather than technical. Ethical and clinical literature increasingly stresses that minimizing false-negatives cannot come at the expense of relational harm or user autonomy.^47,49,52^ Our framework focuses on detection rather than crisis management. Although the proposed *emergency mode* offers a structured response to detected risk, its real-world effectiveness remains to be demonstrated in prospective studies.

In conclusion, mental-health chatbots hold considerable potential to address gaps in access to psychological support, yet recent tragedies have highlighted the fragility of current safety safeguards. Our findings support three interlinked conclusions with implications for the design of safe conversational AI in mental-health context. First, this study supported that suicide- and crisis-risk detection is inherently uncertain. The inter-clinician disagreement and the systematic alignment between model errors and ambiguous cases indicated that crisis risk lacks a definitive ground truth. Therefore, as errors cannot be eliminated, one must move away from performance-based evaluations towards safety-oriented approaches where risk detection is framed as a monitoring problem that explicitly manages uncertaintly in real time. Second, within this uncertainty-aware framing, we show that near–zero–miss risk detection is technically feasible on a clinician-validated dataset. Third, these constraints motivate an architectural rather than purely model-level solution as safety cannot be solved on a classifier level alone. Instead, risk detection must operate as an independent layer, while the conversational system must remain capable of supportive engagement when risk is identified. We describe this principle as an *emergency mode*, described as a state in which the system prioritizes safety without abandoning the user. Together, these findings shift the focus from whether LLMs can recognize crisis toward how systems should respond once risk is detected. By grounding safety mechanisms in uncertainty-aware detection and architectural separation, this work outlines a path toward just-in-time, adaptive safety strategies suitable for real-world mental-health chatbot deployment. As conversational AI increasingly mediates psychological support, layered safety architectures that prioritize harm prevention while preserving user-centered engagement will be essential to ensure that accessibility and empathy do not come at the cost of user safety.^52^

## Declaration Statements

### (1) Dataset Availability

The dataset created for this study is not publicly available due to privacy concerns regarding real patient conversations. The prompts for the models are available upon reasonable request sent to. The code for the different sensitivity levels are available here: https://github.com/sonia-health/llm-mental-health-risk-detection.

## Author Information

### The MULTICAST Consortium

Sebastian Olbrich, MD^1,2^, Birgit Kleim, PhD^1,4^, Katharina Schultebraucks, PhD^9^, Samantha Weber, PhD^1,2*^, Stephanie Homan, PhD^1,4^, Sapir Gershov^9^, Anna Monn, MSc^1,2^, Andrea Häfliger, MSc^1,4^, Jacopo Mocellin, MSc^1,4^

### Author affiliations

1 Psychiatric University Hospital Zurich, Department of Adult Psychiatry and Psychotherapy, Psychiatric University Clinic Zurich, 8032 Zürich, Switzerland

2 Faculty of Medicine, University of Zurich, 8032 Zürich, Switzerland

4 Department of Psychology, University of Zurich, 8052 Zürich, Switzerland.

9 Department of Psychiatry, New York University School of Medicine, New York, NY, USA

## References

1. World Health Organization. Over a billion people living with mental health conditions, services require urgent scale-up. (2025).

2. World Health Organization. Suicide data. (2024).

3. Centers for Disease Control and Prevention. Suicide Rates by Demographic Characteristics, United States, 2003–2023. Morbidity and Mortality Weekly Report (MMWR) 74, 1121–1127 (2024).

4. Ghafari, M., Nadi, T., Bahadivand-Chegini, S. & Doosti-Irani, A. Global prevalence of unmet need for mental health care among adolescents: A systematic review and meta-analysis. Archives of Psychiatric Nursing 36, 1–6 (2022).

5. Zao-Sanders, M. How People Are Really Using Gen AI in 2025. (2025).

6. Sentio University. How Are People Using LLMs for Mental Health Support? (2025).

7. New York Times. OpenAI Sued Over ChatGPT’s Alleged Role in California Teen’s Suicide. (2025).

8. Reuters. OpenAI, Altman Sued Over ChatGPT’s Role in California Teen’s Suicide. (2025).

9. CBS News. Florida mother files lawsuit against AI company over teen son’s death: ‘Addictive and manipulative’. (2024).

10. Wired. ChatGPT, Psychosis, and Self-Harm: What the Data Reveals. (2025).

11. Anthropic. How people use Claude for support, advice, and companionship. (2025).

12. OpenAI. Strengthening ChatGPT’s responses in sensitive conversations. (2025).

13. Dominguez-Olmedo, R., Hardt, M. & Mendler-Dünner, C. Questioning the Survey Responses of Large Language Models. in Advances in Neural Information Processing Systems 37 45850–45878 (Neural Information Processing Systems Foundation, Inc. (NeurIPS), Vancouver, BC, Canada, 2024). doi:10.52202/079017-1458.

14. Campbell, L. O., Babb, K., Lambie, G. W. & Hayes, B. G. An Examination of Generative AI Response to Suicide Inquires: Content Analysis. JMIR Ment Health 12, e73623–e73623 (2025).

15. Haque, M. D. R. & Rubya, S. An Overview of Chatbot-Based Mobile Mental Health Apps: Insights From App Description and User Reviews. JMIR Mhealth Uhealth 11, e44838 (2023).

16. Gould, M. S. et al. National Suicide Prevention Lifeline crisis chat interventions: Evaluation of chatters’ perceptions of effectiveness. Suicide & Life Threat Behav 51, 1126–1137 (2021).

17. Habicht, J. et al. Closing the accessibility gap to mental health treatment with a personalized self-referral chatbot. Nat Med 30, 595–602 (2024).

18. Hawke, L. D. et al. Recommendations for the development of digital conversational agents for youth with mental health conditions: A position paper. DIGITAL HEALTH 11, 20552076251393355 (2025).

19. Li, Z. et al. INSIGHTFUL: insight generation through clinical annotation, analysis and modelling of suicide-related factors towards understanding and lifesaving. BMJ Digit Health 1, e000019 (2025).

20. McBain, R. K. et al. Evaluation of Alignment Between Large Language Models and Expert Clinicians in Suicide Risk Assessment. PS 76, 944–950 (2025).

21. Barak-Corren, Y. et al. Validation of an Electronic Health Record–Based Suicide Risk Prediction Modeling Approach Across Multiple Health Care Systems. JAMA Netw Open 3, e201262 (2020).

22. Levis, M., Leonard Westgate, C., Gui, J., Watts, B. V. & Shiner, B. Natural language processing of clinical mental health notes may add predictive value to existing suicide risk models. Psychol. Med. 51, 1382–1391 (2021).

23. Poulin, C. et al. Predicting the Risk of Suicide by Analyzing the Text of Clinical Notes. PLoS ONE 9, e85733 (2014).

24. De Choudhury, M., Kiciman, E., Dredze, M., Coppersmith, G. & Kumar, M. Discovering Shifts to Suicidal Ideation from Mental Health Content in Social Media. in Proceedings of the 2016 CHI Conference on Human Factors in Computing Systems 2098–2110 (ACM, San Jose California USA, 2016). doi:10.1145/2858036.2858207.

25. Levi-Belz, Y. et al. Predicting imminent suicide risk in a crisis hotline chat using machine learning. Sci Rep 15, 44742 (2025).

26. Kolochowski, F. D., Kreckeler, N., Forkmann, T. & Teismann, T. Reliability of Suicide Risk Estimates: A Vignette Study. Archives of Suicide Research 29, 544–555 (2025).

27. Hashemi, H., Eisner, J., Rosset, C., Van Durme, B. & Kedzie, C. LLM-Rubric: A Multidimensional, Calibrated Approach to Automated Evaluation of Natural Language Texts. in Proceedings of the 62nd Annual Meeting of the Association for Computational Linguistics (Volume 1: Long Papers) 13806–13834 (Association for Computational Linguistics, Bangkok, Thailand, 2024). doi:10.18653/v1/2024.acl-long.745.

28. Lauderdale, S. A. et al. Effectiveness of generative AI-large language models’ recognition of veteran suicide risk: a comparison with human mental health providers using a risk stratification model. Front. Psychiatry 16, 1544951 (2025).

29. Balt, E. et al. Deductively coding psychosocial autopsy interview data using a few-shot learning large language model. Front. Public Health 13, 1512537 (2025).

30. Levkovich, I., et al. Editorial: Empowering suicide prevention efforts with generative AI technology. Front. Psychiatry 16, 1643893 (2025).

31. Boucher, E. M. et al. Artificially intelligent chatbots in digital mental health interventions: a review. Expert Review of Medical Devices 18, 37–49 (2021).

32. Mayor, E. Chatbots and mental health: a scoping review of reviews. Curr Psychol 44, 13619–13640 (2025).

33. Danieli, M., Ciulli, T., Mousavi, S. M. & Riccardi, G. A Conversational Artificial Intelligence Agent for a Mental Health Care App: Evaluation Study of Its Participatory Design. JMIR Form Res 5, e30053 (2021).

34. Melo, A., Silva, I. & Lopes, J. ChatGPT: A Pilot Study on a Promising Tool for Mental Health Support in Psychiatric Inpatient Care. International Journal of Psychiatric Trainees 2, (2024).

35. Joshi, A. C., Ghogare, A. S. & Madavi, P. B. Systematic review of artificial intelligence enabled psychological interventions for depression and anxiety: A comprehensive analysis. Industrial Psychiatry Journal 34, 158–166 (2025).

36. Heinz, M. V., et al. Randomized Trial of a Generative AI Chatbot for Mental Health Treatment. NEJM AI 2, (2025).

37. Gratch, I. & Essig, T. A Letter about “Randomized Trial of a Generative AI Chatbot for Mental Health Treatment”. NEJM AI 2, (2025).

38. Hager, P. et al. Evaluation and mitigation of the limitations of large language models in clinical decision-making. Nat Med 30, 2613–2622 (2024).

39. Thirunavukarasu, A. J. et al. Large language models in medicine. Nat Med 29, 1930–1940 (2023).

40. Riedemann, L., Labonne, M. & Gilbert, S. The path forward for large language models in medicine is open. npj Digit. Med. 7, 339 (2024).

41. Qiu, J., et al. EmoAgent: Assessing and Safeguarding Human-AI Interaction for Mental Health Safety. Preprint at 10.48550/ARXIV.2504.09689 (2025).

42. Rizzo, A., Mozgai, S., Sigaras, A., Rubin, J. E. & Jotwani, R. Expert Consensus Best Practices for the Safe, Ethical, and Effective Design and Implementation of Artificially Intelligent Conversational Agent (i.e., Chatbot/Virtual Human) Systems in Health Care Applications. Journal of Medical Extended Reality 2, 209–222 (2025).

43. Nahum-Shani, I. et al. Just-in-Time Adaptive Interventions (JITAIs) in Mobile Health: Key Components and Design Principles for Ongoing Health Behavior Support. Annals of Behavioral Medicine 52, 446–462 (2018).

44. Coppersmith, D. D. L. et al. Just-in-Time Adaptive Interventions for Suicide Prevention: Promise, Challenges, and Future Directions. Psychiatry 85, 317–333 (2022).

45. The Guardian. Has OpenAI really made ChatGPT better for users with mental health problems? (2025).

46. Zortea, T. C., Cleare, S., Melson, A. J., Wetherall, K. & O’Connor, R. C. Understanding and managing suicide risk. British Medical Bulletin 134, 73–84 (2020).

47. Gould, M. S. et al. Helping Callers to the National Suicide Prevention Lifeline Who Are at Imminent Risk of Suicide: Evaluation of Caller Risk Profiles and Interventions Implemented. Suicide & Life Threat Behav 46, 172–190 (2016).

48. Gould, M. S. et al. National Suicide Prevention Lifeline (Now 988 Suicide and Crisis Lifeline): Evaluation of Crisis Call Outcomes for Suicidal Callers. Suicide & Life Threat Behav 55, e70020 (2025).

49. Ryan, E. P. & Oquendo, M. A. Suicide Risk Assessment and Prevention: Challenges and Opportunities. FOC 18, 88–99 (2020).

50. Ward-Ciesielski, E. F. & Rizvi, S. L. The potential iatrogenic effects of psychiatric hospitalization for suicidal behavior: A critical review and recommendations for research. Clinical Psychology: Science and Practice 28, 60–71 (2021).

51. Banks, J. Deletion, departure, death: Experiences of AI companion loss. Journal of Social and Personal Relationships 41, 3547–3572 (2024).

52. Goldberg, C. B. et al. To do no harm — and the most good — with AI in health care. Nat Med 30, 623–627 (2024).

